# External validation of risk scores to predict in-hospital mortality in patients hospitalized due to coronavirus disease 2019

**DOI:** 10.1101/2022.03.11.22271912

**Authors:** Shermarke Hassan, Chava L. Ramspek, Barbara Ferrari, Merel van Diepen, Raffaella Rossio, Rachel Knevel, Vincenzo la Mura, Andrea Artoni, Ida Martinelli, Alessandra Bandera, Alessandro Nobili, Andrea Gori, Francesco Blasi, Ciro Canetta, Nicola Montano, Frits R. Rosendaal, Flora Peyvandi, LUMC-COVID-19 Research Group, COVID-19 Network working group

## Abstract

**Background:** The coronavirus disease 2019 (COVID-19) presents an urgent threat to global health. Prediction models that accurately estimate mortality risk in hospitalized patients could assist medical staff in treatment and allocating limited resources.

**Aims:** To externally validate two promising previously published risk scores that predict in-hospital mortality among hospitalized COVID-19 patients.

**Methods:** Two cohorts were available; a cohort of 1028 patients admitted to one of nine hospitals in Lombardy, Italy (the Lombardy cohort) and a cohort of 432 patients admitted to a hospital in Leiden, the Netherlands (the Leiden cohort). The primary endpoint was in-hospital mortality. All patients were adult and tested COVID-19 PCR-positive. Model discrimination and calibration were assessed.

**Results:** The C-statistic of the 4C mortality score was good in the Lombardy cohort (0.85, 95CI: 0.82-0.89) and in the Leiden cohort (0.87, 95CI: 0.80-0.94). Model calibration was acceptable in the Lombardy cohort but poor in the Leiden cohort due to the model systematically overpredicting the mortality risk for all patients. The C-statistic of the CURB-65 score was good in the Lombardy cohort (0.80, 95CI: 0.75-0.85) and in the Leiden cohort (0.82, 95CI: 0.76-0.88). The mortality rate in the CURB-65 development cohort was much lower than the mortality rate in the Lombardy cohort. A similar but less pronounced trend was found for patients in the Leiden cohort.

**Conclusion:** Although performances did not differ greatly, the 4C mortality score showed the best performance. However, because of quickly changing circumstances, model recalibration may be necessary before using the 4C mortality score.

## Introduction

The coronavirus disease 2019 (COVID-19) epidemic, which started in early December 2019, presents an important threat to global health. As of October 18^th^ 2021, the number of patients confirmed to have the disease has exceeded 240 million and more than 4,900,537 people have died from COVID-19 infection. [1]

The outbreak overwhelmed the healthcare system in several countries, leading to shortages in hospital beds and medical equipment. [2–4] Prediction models that estimate the risk of hospitalized patients experiencing a poor outcome could assist medical staff in triaging patients when allocating limited healthcare resources.

A systematic review of prognostic prediction models for poor outcomes in hospitalized COVID-19 patients [5] reported that most models showed good predictive performance, but almost all models had a high risk of bias owing to a combination of poor reporting and poor methodological conduct for participant selection, predictor description, and/or statistical methods used. Of the 107 reviewed prognostic scores, only one, the 4C mortality score [6], was identified as being of good methodological quality. [5] The 4C mortality score development cohort consisted of 35,463 patients enrolled between February 6^th^ 2020 and May 20^th^ 2020. Several external validation studies reported C-statistics between 0.78 and 0.84. [7–11]

The systematic review by Wynants et al. [5] also identified another prediction model of interest [12]. In addition to reporting on the quality of newly developed COVID-19 specific prediction models, the review also reported on several previously published “general-purpose” mortality risk prediction models that were externally validated in the COVID-19 population. Among these models, the CURB-65 score [13], was also found to be a promising candidate model. The CURB-65 development cohort consisted of 718 patients with community-acquired pneumonia enrolled between October 1998 and December 2000. The CURB-65 score has been externally validated in COVID-19 patients in several studies that reported C-statistics ranging from 0.58-84. [12, 14–17] The CURB-65 score was also directly compared with the 4C mortality score. This was done in the external validation cohort (N = 15,560) used in the same publication that also reported on the development of the 4C mortality score. The C-statistic was 0.72 (0.71-0.73). [6]

Unfortunately, few studies have properly evaluated the 4C mortality score and the CURB-65. Many studies lacked the sample size to properly externally validate these scores. Furthermore, most studies only assessed model discrimination (by calculating a C-statistic) but not model calibration (which refers to the degree to which the mortality risk predicted by a given model is in agreement with the observed risk). In addition, none of these studies considered recalibrating the 4C mortality score to increase the performance of this score in the local population. Poorly calibrated models can lead to wrong clinical decisions, as the predicted mortality risk for a given patient may be widely different from the actual mortality risk. [18] Therefore, the aim of the study was to externally validate two promising risk tools that predict mortality among hospitalized COVID-19 positive patients; one COVID-19-specific score (the 4C mortality score) and one general-purpose score (the CURB-65 score) in a cohort of 1028 COVID-19 positive patients admitted to one of nine hospitals in Lombardy, Italy (the Lombardy cohort) and a cohort of 432 COVID-19 positive patients admitted to a hospital in Leiden, the Netherlands (the Leiden cohort).

## Methods

### Study design

Two cohorts of adult patients diagnosed with COVID-19 were available for external validation. The first cohort consisted of adult patients hospitalized at one of nine hospitals in the province of Lombardy, Italy (the Lombardy cohort). The second cohort consisted of adult patients hospitalized at Leiden University Medical Center, Leiden, the Netherlands (the Leiden cohort). Patients were included at hospital admission. For both cohorts, patients that were transferred from other hospitals, and patients that were directly admitted to the ICU were excluded. This study was approved by the Medical Ethics Committee of the Fondazione IRCCS Ca’ Granda Ospedale Maggiore Policlinico and the Institutional Review Board of the LUMC for observational studies.

### Data collection

Clinical data were collected using a case report form (which was based on the ISARIC-WHO case report form). Only data that were available during the first 24 hours of admission were used. If multiple values were present, the earliest recorded value was used.

#### Predictor variables for the 4C mortality score

The 4C mortality score was developed in a cohort of COVID-19 PCR-positive adult patients that were admitted to one of 260 hospitals in England, Scotland, or Wales between 6 February and 20 May 2020. The outcome was in-hospital mortality. The 4C mortality score includes the following eight predictors, collected on the day of admission: age in years (categorical variable: <50, 50-59, 60-69, 70-79, ≥80); sex at birth (dichotomous variable: male, female); respiratory rate in breaths/min (categorical variable: >20, 20-29, ≥30); oxygen saturation on room air (dichotomous variable: ≥92%, <92%); Glasgow coma scale (dichotomous variable: 15 points, <15 points); urea (categorical variable: <7 mmol/L, ≥7 to ≤14 mmol/L, >14 mmol/L); CRP (categorical variable: <50 mg/L, 50-99 mg/L, ≥100 mg/L) and the number of comorbidities. The list of comorbidities is based on the Charlson comorbidity index [19] with the addition of clinician-defined obesity.

#### Predictor variables for the CURB-65 score

The CURB-65 score was developed in a cohort of adult patients admitted as medical emergencies with community acquired pneumonia (CAP) to hospitals in the UK, New Zealand, and the Netherlands between October 1998 and December 2000. The outcome was 30-day mortality. The CURB-65 score consisted of the following 5 predictors that were to be measured at the emergency department: mental confusion, defined as a score of ≤ 8 on the Abbreviated Mental Test score; urea (categorical variable: ≤7 mmol/L, >7 mmol/L); respiratory rate in breaths/min (categorical variable: <30, ≥30); a systolic blood pressure of < 90 mmHg or a diastolic blood pressure of ≤ 60 mmHg (dichotomous variable; no, yes) and age (dichotomous variable; <65 years, ≥65 years). If information on mental confusion was missing, a Glasgow Coma Scale score of ≤ 14 was used as an indicator of mental confusion. (this method was also used previous studies [20])

#### Outcome

The outcome used in this study for all models was 30-day in-hospital mortality.

### External validation of all risk scores

For the 4C mortality score, the predicted risk of 30-day in-hospital mortality was calculated for each individual in the validation cohort. The model formula and coefficients for the 4C mortality score were obtained from the supplement of the original paper. [6] In addition, the number of points that each individual scored on the 4C mortality score was also calculated. The original publication of the CURB-65 score did not provide the model formula underlying the score, only the score itself. [13] Therefore, it was not possible to calculate the individual predicted risk. Instead, only the individual CURB-65 score was calculated for each patient.

Model discrimination for all models was assessed using the C-statistic. The C-statistic reflects the degree to which a model can distinguish between patients with and patients without the outcome and ranges from 0.5 (no discrimination) to 1.0 (perfect discrimination). [21] For the 4C mortality score, the C-statistic was calculated in two ways; via the predicted risk of 30-day in-hospital mortality (which was calculated from the regression model underlying the risk tool), and via the simplified points score (which was constructed to facilitate usage of the risk tool by clinicians). For the CURB-65, only the points score was available and we therefore only used the points score to calculate model discrimination.

Model calibration of the 4C mortality score was assessed visually by plotting calibration curves. [21] To construct the calibration curve, the predicted outcome probabilities were plotted against observed outcome frequencies, for each quintile of predicted risk. To examine calibration across the whole range, a LOESS (Locally Estimated Scatterplot Smoothing) line was estimated. As it was not possible to obtain the individual predicted mortality risk using the CURB-65 score, plotting a calibration curve was not possible. Instead, an alternative method, also implemented in other studies [22, 23], was used. Patients were divided into groups, based on the number of points on the CURB-65 score. Next, a bar chart was constructed where the proportion of deaths of each group in the external validation cohort was compared against the proportion of deaths in the same group in the CURB-65 development cohort.

The 4C mortality score was also recalibrated to better reflect the mortality incidence in the validation cohorts. Recalibration methods range from conservative to very extensive. Two recalibration methods were used. The first recalibration method was the most conservative and consisted of fitting a logistic regression model to the validation cohort dataset, with the intercept as a free parameter and the linear predictor of the 4C mortality score as an offset variable. [24] This method corrects for systematic over- or underprediction by the model. The second recalibration method was more extensive and consisted of fitting the same logistic regression model as described above, but leaving both the intercept and the coefficient for the linear predictor to be freely estimated. [24] A more detailed explanation of this methodology, as well as the details of the various recalibrated models presented in the results section, are given in the Appendix. As the predicted mortality risk for each patient was not available for the CURB-65 score, model recalibration was not done for this score.

### Handling missing values

Missing values in the Lombardy cohort and Leiden cohort datasets were imputed using multivariate imputation by chained equations. [25] The C-statistic was pooled using Rubin’s rules. [26] All imputed datasets were combined to create one dataset, which was then used to create the calibration plots.

### Statistical packages

All analyses were performed using R version 3.6.2.

### Data sharing

The datasets generated during and/or analyzed during the current study are available from the corresponding author on reasonable request.

## Results

### General characteristics

The Lombardy cohort consisted of 1028 patients enrolled between February 25^th^ 2020 and August 1^st^ 2020. The Leiden cohort consisted of 432 patients enrolled between March 7^th^ 2020 and March 5^th^ 2021.

The mortality rate was 21% in the Lombardy cohort, 10% in the Leiden cohort, 32% in the 4C mortality score development cohort and 10% in the CURB-65 score development cohort. (Table 1) The median age for all patients was 66 years in the Lombardy cohort, 65 years in the Leiden cohort, 73 years in the 4C mortality score development cohort. (Table 1) The portion of patients with ≥ 2 comorbidities was 24% in the Lombardy cohort, 35% in the Leiden cohort and 48% in the 4C mortality score development cohort. (Table 1)

**Table 1:**
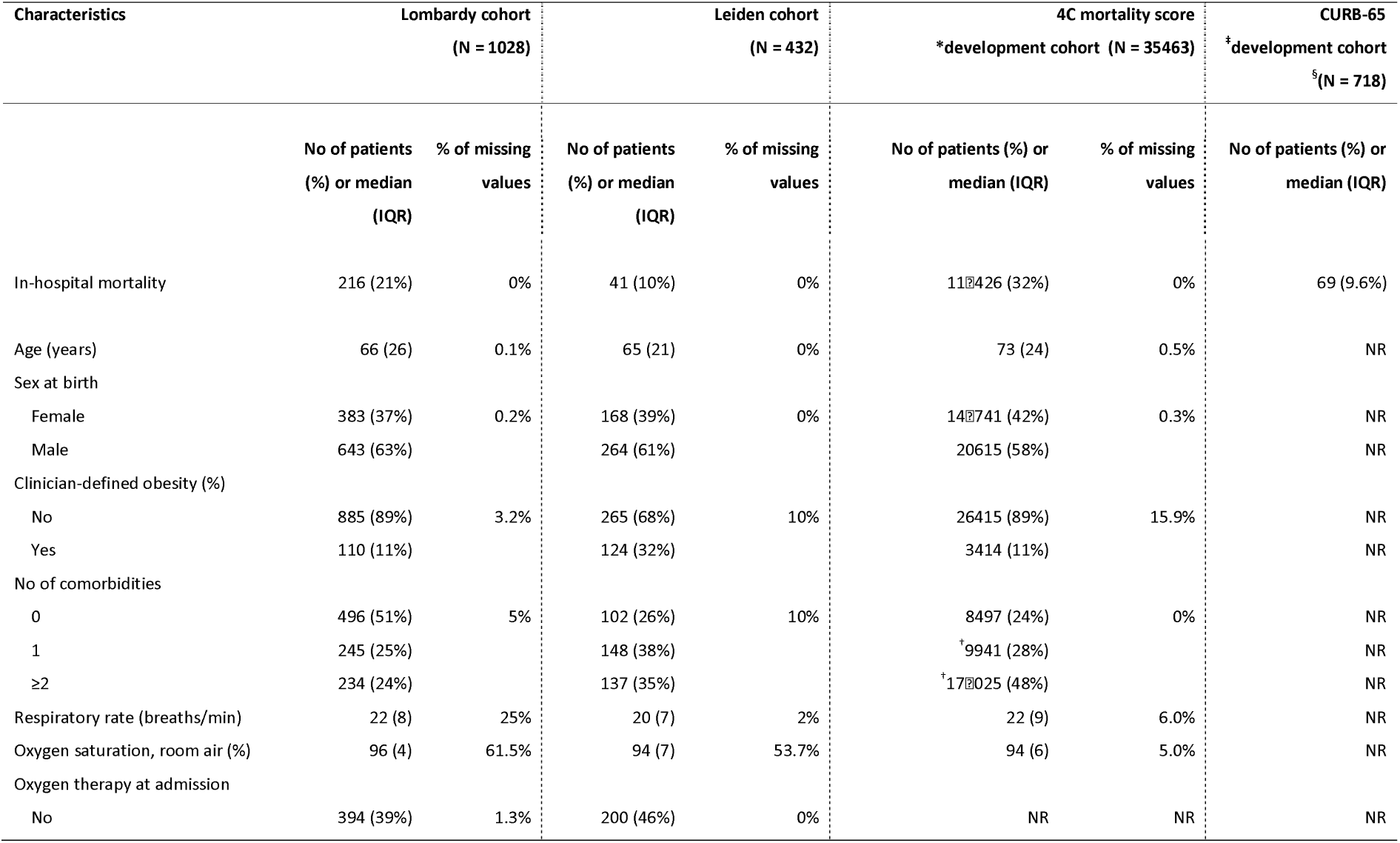
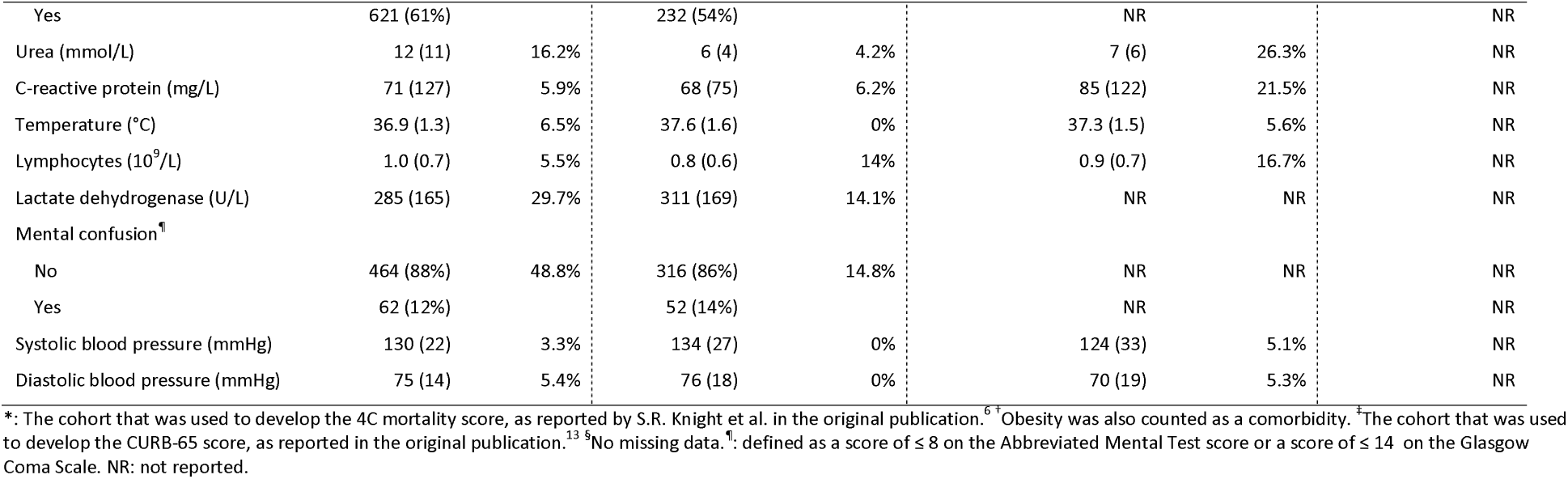
General characteristics

### External validation of 4C mortality score

The C-statistic of the 4C mortality score in the Lombardy cohort, which was calculated based on the predicted risk of 30-day in-hospital mortality obtained from the underlying regression model, was 0.85 (95CI: 0.82-0.89). (Table 2, Figure 1) The C-statistic of the 4C mortality score in the Lombardy cohort, calculated based on the simplified points score, was exactly the same (C-statistic: 0.85, 95CI: 0.82-0.89). Calibration was acceptable. However, the 4C mortality score did overpredict the risk of mortality in the 0-50% risk range. (Figure 2A) After applying the first recalibration method, the 4C mortality score showed better calibration in the lower risk range but underpredicted the risk in the >50% risk range. (Figure 2B) Applying the second recalibration method resulted in almost perfect calibration. (Figure 2C)

**Table 2:** C-statistic of the 4C mortality score and the CURB-65 score during external validation

| Models | Lombardy cohort | Leiden cohort |
| --- | --- | --- |
| 4C mortality score | 0.85 (95CI: 0.82-0.89) | 0.87 (95CI: 0.80-0.94) |
| CURB-65 score | 0.80 (95CI: 0.75-0.85) | 0.82 (95CI: 0.76-0.88) |

\*: Not applicable, model was developed in this cohort.

**Figure 1:**
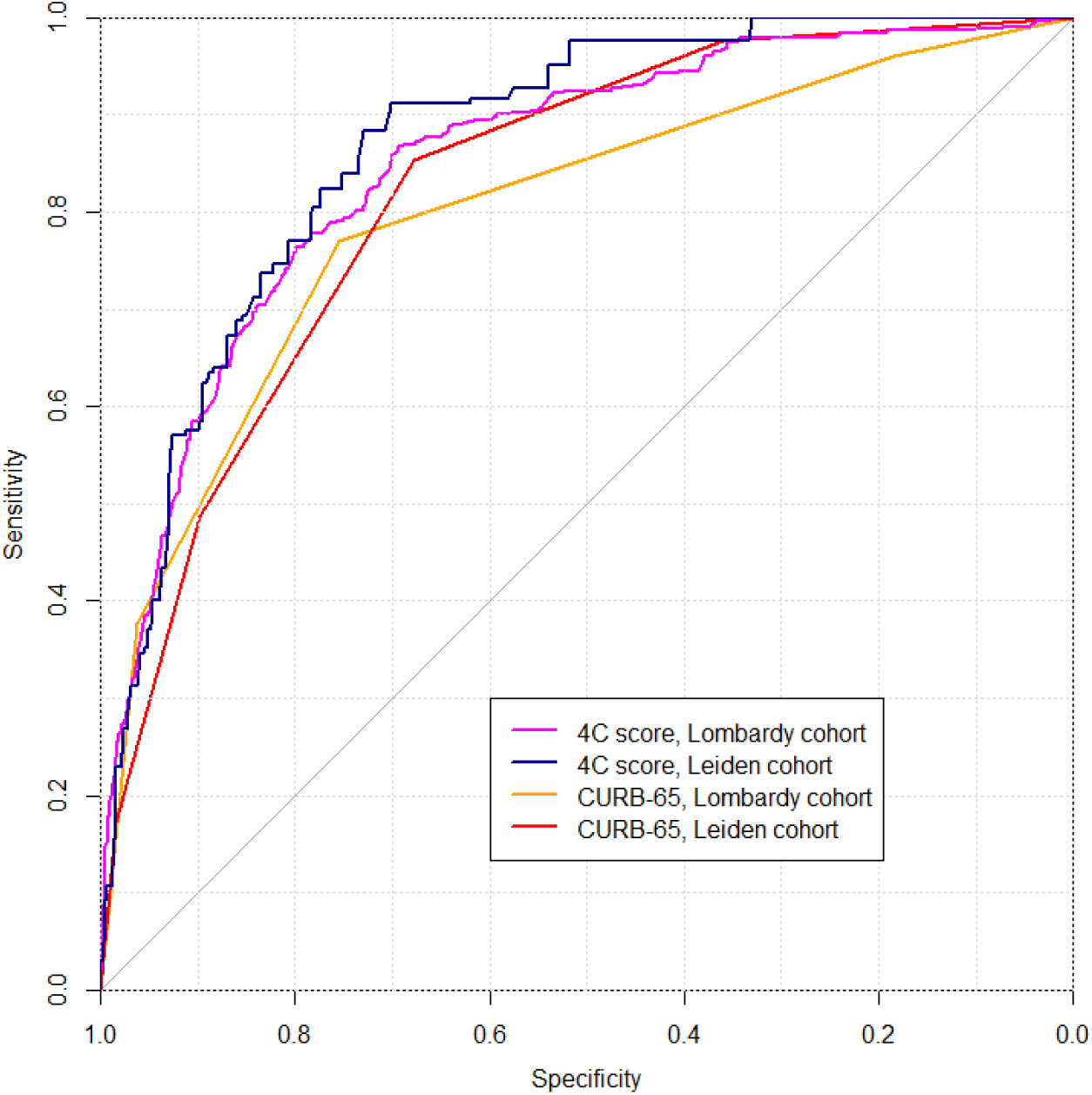
Receiver operating characteristic (ROC) curve The figure shows the ROC curve for the 4C mortality score and the CURB-65 score for both the Lombardy cohort and the Leiden cohort. The area under the curve is equal to the C-statistic, and varies from 0.5 (poor discrimination, basically the same as flipping a coin to determine the outcome) to 1 (the model can discriminate perfectly between patients that died and patients that survived). The C-statistics and corresponding confidence intervals are reported in Table 2.

**Figure 2:**
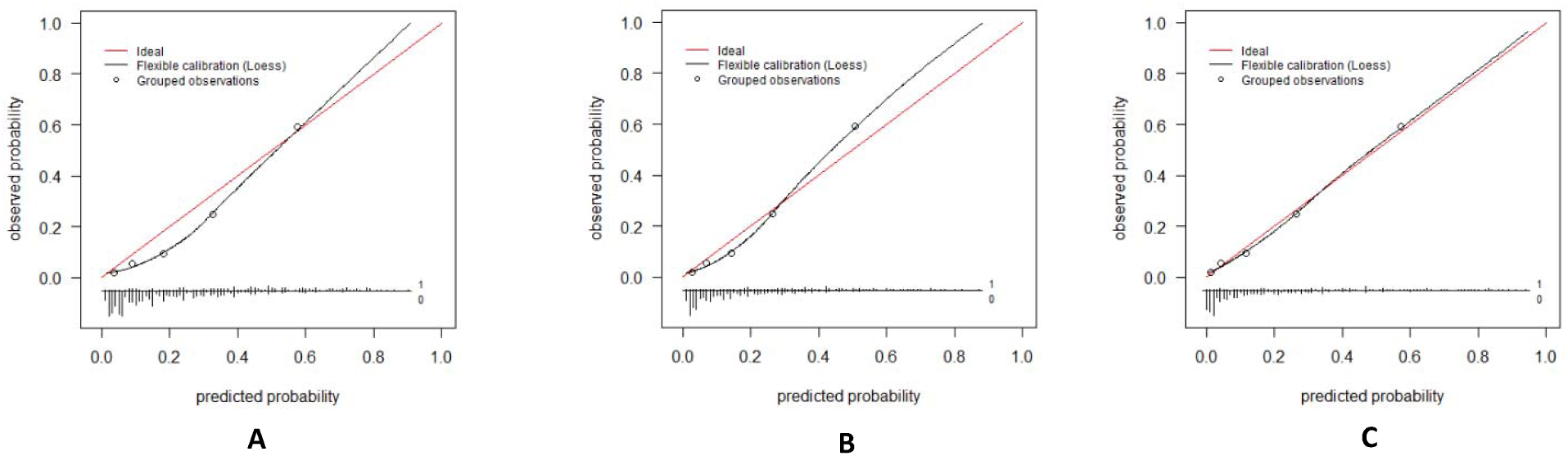
Calibration of the standard 4C mortality score and the recalibrated models in the Lombardy cohort Model calibration was assessed visually by plotting calibration curves. Figure **2A** represents the calibration plot for the standard model in the Lombardy cohort. Figure **2B** represents the calibration plot for the first recalibrated model in the Lombardy cohort. (only intercept was re-estimated) Figure **2C** represents the calibration plot for the second recalibrated model in the Lombardy cohort. (intercept and slope were re-estimated) To construct the calibration curve, the cohort was divided into quintiles based on the predicted mortality risk. Next, the predicted mortality risk of each group was plotted against the observed mortality rate for that group. (these points are represented as circles) Ideally, the predicted and observed mortality risk should be equal, for each group. With ideal model calibration, all points should fall on the diagonal red line. To examine model calibration across the entire risk range, a LOESS (Locally Estimated Scatterplot Smoothing) line was estimated. Below the calibration curves are histograms showing the distribution of predicted probabilities for patients that died during follow-up (all bars above the horizontal line, labeled ‘1’) and for patients that survived (all bars below the horizontal line, labeled ‘0’). The length of a bar corresponds to the number of patients with that predicted probability.

The C-statistic of the 4C mortality score in the Leiden cohort, which was calculated based on the predicted risk of 30-day in-hospital mortality obtained from the underlying regression model, was 0.87 (95CI: 0.80-0.94). (Table 2, Figure 1) The C-statistic of the 4C mortality score in the Leiden cohort, calculated based on the simplified points score, was exactly the same (C-statistic: 0.87, 95CI: 0.80-0.94). Calibration was poor as the 4C mortality score overpredicted the mortality rate across the entire risk range. (Figure 3A) After applying the first recalibration method, the 4C mortality score showed slightly improved calibration but now underpredicted the risk across most of the risk range. (Figure 3B) Applying the second recalibration method resulted in a model with very good calibration, which was almost perfect for the 0-50% predicted risk range. (Figure 3C) Calibration in the higher risk ranges was less accurate, but few patients had a predicted risk higher than 50%.

**Figure 3:**
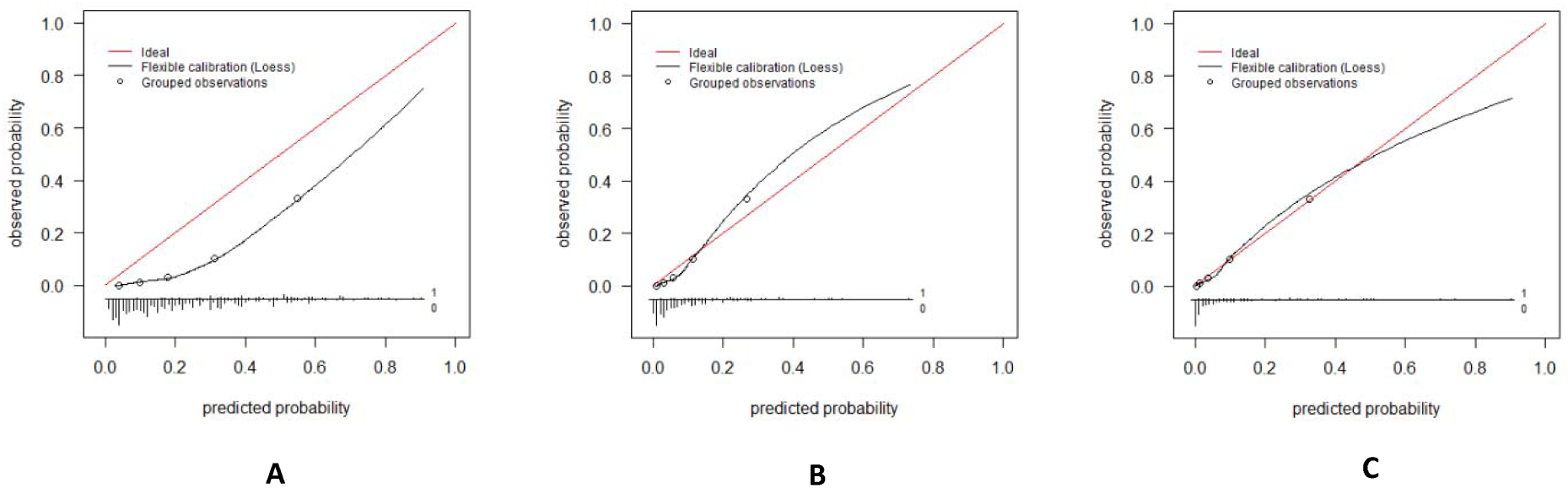
Calibration of the standard 4C mortality score and the recalibrated models in the Leiden cohort Model calibration was assessed visually by plotting calibration curves. Figure **3A** represents the calibration plot for the standard model in the Leiden cohort. Figure **3B** represents the calibration plot for the first recalibrated model in the Leiden cohort. (only intercept was re-estimated) Figure **3C** represents the calibration plot for the second recalibrated model in the Leiden cohort. (intercept and slope were re-estimated) To construct the calibration curve, the cohort was divided into quintiles based on the predicted mortality risk. Next, the predicted mortality risk of each group was plotted against the observed mortality rate for that group. (these points are represented as circles) Ideally, the predicted and observed mortality risk should be equal, for each group. With ideal model calibration, all points should fall on the diagonal red line. To examine model calibration across the entire risk range, a LOESS (Locally Estimated Scatterplot Smoothing) line was estimated. Below the calibration curves are histograms showing the distribution of predicted probabilities for patients that died during follow-up (all bars above the horizontal line, labeled ‘1’) and for patients that survived (all bars below the horizontal line, labeled ‘0’). The length of a bar corresponds to the number of patients with that predicted probability.

### External validation of CURB-65 score

The C-statistic of the CURB-65 score in the Lombardy cohort was 0.80 (95CI: 0.75-0.85). (Table 2, Figure 1) The observed mortality risk for patients in the CURB-65 development cohort was much lower than the observed mortality risk for patients in the Leiden cohort, for every group of patients with a specific CURB-65 score. (Figure 4) The C-statistic of the CURB-65 score in the Leiden cohort was 0.82 (95CI: 0.76-0.88). (Table 2, Figure 1) The observed mortality risk for patients in the CURB-65 development cohort was slightly lower than the observed mortality risk for patients in the Leiden cohort, especially for patients with ≥ 2 points on the CURB-65 score. (Figure 4)

**Figure 4:**
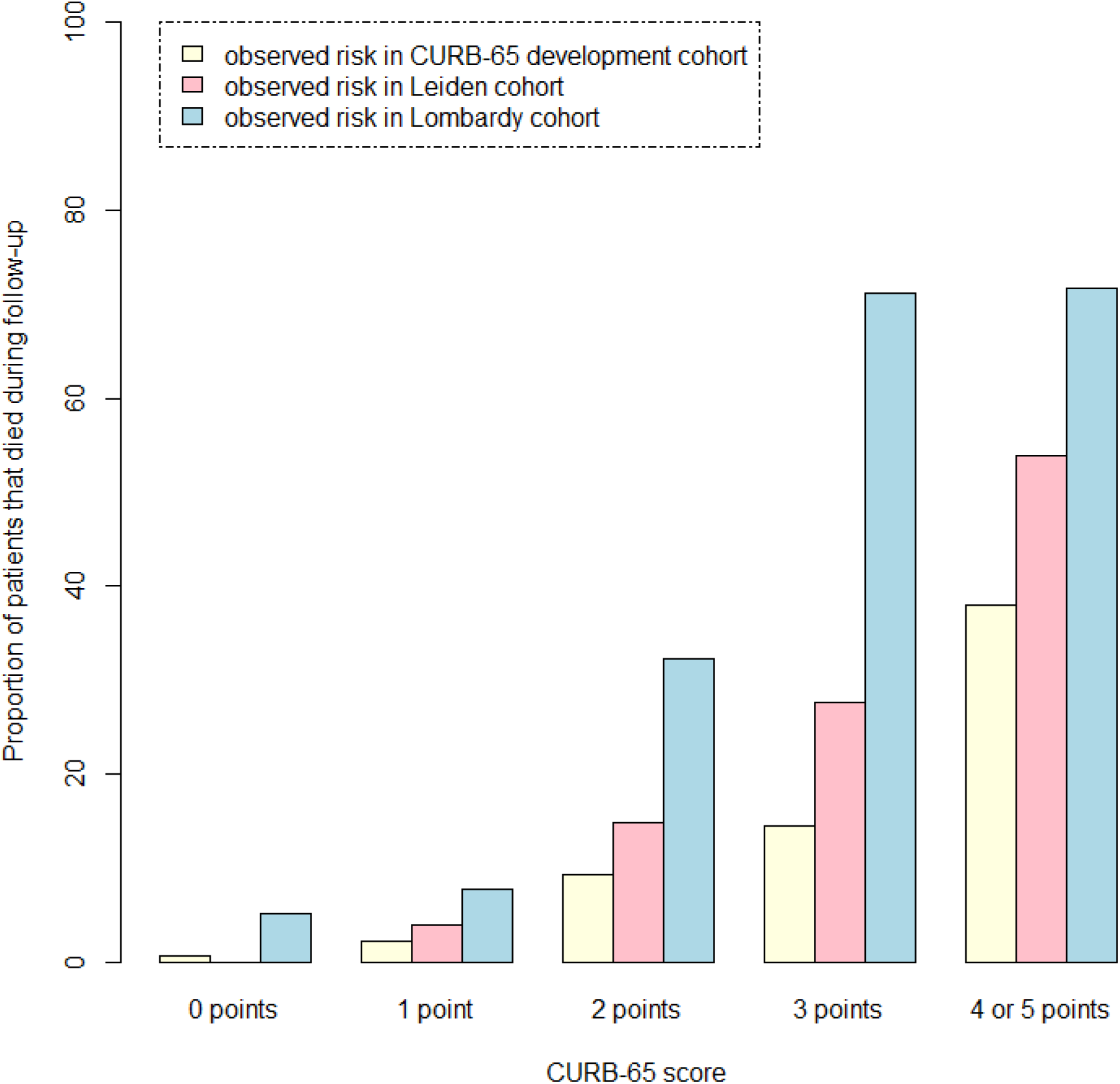
Calibration of the CURB-65 score in the Lombardy cohort and Leiden cohort Model calibration was assessed visually in bar chart. Each cohort was divided into groups, based on the number of points on the CURB-65 score. For each group, the observed proportion of deaths in the CURB-65 development cohort (represented by the light-yellow bars) was compared against the observed proportion of deaths in the Leiden cohort (represented by the pink bars) and the Lombardy cohort (represented by the light-blue bars). Ideally, all bars for a given group should have the same height.

## Discussion

### Main findings

Two promising prognostic scores that are used to predict in-hospital mortality in patients hospitalized with COVID-19 were selected for external validation based on the results of a previous systematic review. [5]

Although both models showed good discrimination, the 4C mortality score performed the best, both in the Lombardy cohort (C-statistic: 0.85) as well as the Leiden cohort (C-statistic: 0.87). The 4C model performed less well in terms of model calibration, as the standard model overpredicted the risk in most patients, in both the Lombardy cohort and the Leiden cohort. In the Lombardy cohort, the degree of overprediction was still acceptable but it was unacceptably high in the Leiden cohort. Updating the 4C mortality score to the local setting by applying a conservative recalibration method strongly improved model calibration in the Leiden cohort in the 0-30% risk range (which contained most of the patients in the cohort). Excellent calibration in both cohorts was obtained by applying a second, slightly more extensive, recalibration method.

The CURB-65 score also showed good discrimination. However, direct assessment of model calibration was not possible. We indirectly assessed model calibration by comparing the mortality rate in the development cohort with the mortality rate in the external validation cohort.

The differences in model performance between cohorts can be explained by two factors. [27] Firstly, part of the reduction in model performance is due to overfitting. Secondly, differences in the type of patients admitted to the hospital as well as differences in treatment protocol between the development cohorts and the external validation cohorts could also have influenced model performance.

For example, almost all patients (91%) in the Lombardy cohort were enrolled during the first two months of the first COVID-19 wave in Italy (March and April 2020) while patients in the Leiden cohort were uniformly enrolled from March 7^th^ 2020 and March 5^th^ 2021. This means that most patients in the Leiden cohort would have had access to better treatment, as much more was known about the effectiveness of different COVID-19 treatment options. This will have impacted the performance of the model in the external validation cohort.

Another issue is that the 4C mortality score development cohort was older (73 years) than the Leiden cohort (65 years) and the Lombardy cohort (66 years). This also explains the difference in mortality rate, which was considerably higher in the 4C mortality score development cohort (32.2%) than in the Leiden cohort (8.3%) or the Lombardy cohort (21.7%). Due to this, it was to be expected that the 4C mortality score would systematically overpredict the mortality risk in the external validation cohorts, given that it was developed in a cohort of patients with a much higher average mortality rate. Recalibrating was an adequate way to solve this issue.

### Comparison with other studies

The development of the 4C mortality score was in line with the most recent guidelines for prediction model development. [28] Furthermore, the patient cohorts used to develop and validate the 4C mortality score were extremely large, minimizing the chance of overfitting. The model has since been validated in a number of cohorts from other populations. In a large cohort of 14,343 patients from hospitals in the greater Paris area, the 4C mortality score had a C-statistic of 0.79 (95% CI: 0.78–0.80). Calibration was acceptable, although the model somewhat overpredicted the risk in most patients. [29] in a cohort of 925 Brazilian and 438 Spanish patients, a C-statistic of 0.78 (95%CI: 0.75-0.81) was found. Overall calibration was good in this cohort. [30] Furthermore, the 4C mortality score had a C-statistic of 0.78 (0.70–0.85) in a cohort of 1027 Canadian patients from Toronto. Calibration was not assessed formally but by plotting model scores against observed probabilities, which makes calibration difficult to interpret. [11] Lastly, a Japanese study of 693 patients reported good discrimination (0.84, 95%CI: 0.80-0.88) and calibration. [31] None of the aforementioned studies recalibrated their models to better fit the local population.

The CURB-65 score has also been previously validated in different populations of COVID-19 positive patients. The CURB-65 showed a C-statistic of 0.83 (0.82–0.84) in a very large cohort of 10,328 patients from Spain. [32] Furthermore, a preprint manuscript reported that the CURB-65 score was tested in patients hospitalized in one of thirteen acute care hospitals in the New York City area. The score had a C-statistic of 0.80 in cohort of 2229 patients and 0.72 in another cohort of 3328 patients. [17] In a cohort of 1717 COVID-19 positive patients admitted to a hospital in Shanghai, China, the CURB-65 score had a fairly low C-statistic of 0.70 (95CI: 0.66-0.73). [33] Lastly, in a cohort of 1181 patients from Qatar, a C-statistic of 0.78 (95% CI 0.70-0.86) was reported. [34] Overall, the discriminative performance of the CURB-65 reported by these studies varied from moderate to good. Similar to our study, none of the aforementioned studies assessed model calibration of the CURB-65 score. (as this was not possible)

### Limitations

It has been suggested that the minimum sample size for external validation should be at least 100 events and 100 non-events. [35] The Leiden cohort had 41 deaths, falling below the number suggested by this rule of thumb.

The sample size of the Lombardy cohort was acceptable for external validation. However, the Lombardy cohort consisted of patients that were enrolled in the first months of 2020. After this period, the incidence COVID-19 related mortality has changed a lot due to many treatment changes. This limits the applicability of the recalibrated 4C risk score as the population in which the scores were recalibrated may not be representative of the current patient population in 2022.

Furthermore, some patients who were already very ill before being hospitalized for COVID-19 would have chosen to receive end-of-life care at home. Despite not dying in the hospital (in-hospital mortality was the study outcome), the 4C mortality score would have assigned these patients a very high predicted in-hospital mortality risk. This will have reduced the model performance (especially model discrimination), depending on the proportion of patients that received end-of-life care at home.

Lastly, missing data may have influenced model performance. For example, the oxygen saturation on room air (a predictor in the 4C mortality score) was missing in more than half of all patients in both the Lombardy cohort and the Leiden cohort. This is most likely because these patients were already receiving oxygen therapy at admission.

### Clinical use

Given the gradual uptake of the COVID-19 vaccine, the chances of the COVID-19 virus overburdening hospital resources at the national level in developed countries is becoming smaller, although localized outbreaks might still occur, especially in places with high rates of vaccine hesitancy. On the other hand, vaccine uptake in developing countries is still extremely low [1] and a viral outbreak could severely strain local resources. Furthermore, viral evolution could lead to a novel variant that current vaccines are not or only partially effective against. [36]

Risk scores could be used to identify patients with a high mortality risk. These patients could be candidates for early escalation to critical care while low-risk patients could be safely managed outside the hospital. For this purpose, an accurate estimate of the absolute risk of mortality for a given patient is essential. A risk score is sometimes used in a patient population that is very different (in terms of patient- and treatment characteristics) than the patient population in which the risk score was originally developed. In this situation, it is highly likely that the score will be miscalibrated and model recalibration might be necessary before this score can be used to obtain an accurate estimate of the absolute risk of mortality.

If recalibration of the risk tool is necessary, we would suggest using a more conservative recalibration method that changes the original model as little as possible to minimize overfitting the model to the new setting. Depending on the sample size, more extensive recalibration models can be considered. Recalibrating a model before use might be too complicated for end-users (i.e. clinicians). An app or web tool that automatically produces an adjusted risk score based on a user-specified mean mortality rate would simplify the process significantly and increase usage of these scores among clinicians. Lastly, as vaccination rates increase, an updated model that also includes information on vaccination status might yield better predictions.

In situations where hospital ICUs are overburdened, a risk score could be used to only admit patients with a low predicted mortality risk and transfer high-risk patients to other centers with more ICU capacity. For this purpose, the clinician only needs to know if a given patient has a lower or higher risk of mortality relative to other patients. In this situation, a risk score only needs to have good model discrimination. The 4C mortality score showed good model discrimination (as measured with the C-statistic) across different populations, and could therefore be used in clinical practice for this purpose, without recalibrating the model.

### Conclusion

Two previously published risk scores were externally validated in two different settings. Although performances did not differ greatly, the 4C mortality score showed the best model performance. However, if the reason for using this risk tool is to obtain accurate absolute mortality risks, recalibration of the model to the local patient population might be necessary before use.

## Authorship

### Author contributions

S. Hassan analyzed the data and wrote the manuscript. C.L. Ramspek, M. van Diepen, F. Peyvandi and F.R. Rosendaal interpreted the data and reviewed the manuscript. All other authors were involved in either coordination, patient enrollment, data collection and/or reviewing the manuscript.

## Acknowledgments

We are indebted to all the patients with COVID-19 who participated in this research. We would also like to thank the COVID-19 Network working group and the LUMC-COVID-19 Research Group members. Their names are listed below:

Silvano Bosari, Luigia Scudeller, Giuliana Fusetti, Laura Rusconi, Silvia Dell’Orto, Daniele Prati, Luca Valenti, Silvia Giovannelli, Maria Manunta, Giuseppe Lamorte, Francesca Ferarri, Andrea Gori, Alessandra Bandera, Antonio Muscatello, Davide Mangioni, Laura Alagna, Giorgio Bozzi, Andrea Lombardi, Riccardo Ungaro, Giuseppe Ancona, Gianluca Zuglian, Matteo Bolis, Nathalie Iannotti, Serena Ludovisi, Agnese Comelli, Giulia Renisi, Simona Biscarini, Valeria Castelli, Emanuele Palomba, Marco Fava, Valeria Fortina, Carlo Alberto Peri, Paola Saltini, Giulia Viero, Teresa Itri, Valentina Ferroni, Valeria Pastore, Roberta Massafra, Arianna Liparoti, Toussaint Muheberimana, Alessandro Giommi, Rosaria Bianco, Rafaela Montalvao De Azevedo, Grazia Eliana Chitani, Flora Peyvandi, Roberta Gualtierotti, Barbara Ferrari, Raffaella Rossio, Nadia Boasi, Erica Pagliaro, Costanza Massimo, Michele De Caro, Andrea Giachi, Nicola Montano, Barbara Vigone, Chiara Bellocchi, Angelica Carandina, Elisa Fiorelli, Valerie Melli, Eleonora Tobaldini, Francesco Blasi, Stefano Aliberti, Maura Spotti, Leonardo Terranova, Sofia Misuraca, Alice D’Adda, Silvia Della Fiore, Marta Di Pasquale, Marco Mantero, Martina Contarini, Margherita Ori, Letizia Morlacchi, Valeria Rossetti, Andrea Gramegna, Maria Pappalettera, Mirta Cavallini, Agata Buscemi, Marco Vicenzi, Irena Rota, Giorgio Costantino, Monica Solbiati, Ludovico Furlan, Marta Mancarella, Giulia Colombo, Giorgio Colombo, Alice Fanin, Mariele Passarella, Valter Monzani, Ciro Canetta, Angelo Rovellini, Laura Barbetta, Filippo Billi, Christian Folli, Silvia Accordino, Diletta Maira, Cinzia Maria Hu, Irene Motta, Natalia Scaramellini, Anna Ludovica Fracanzani, Rosa Lombardi, Annalisa Cespiati, Matteo Cesari, Tiziano Lucchi, Marco Proietti, Laura Calcaterra, Clara Mandelli, Carlotta Coppola, Arturo Cerizza, Antonio Maria Pesenti, Giacomo Grasselli, Alessandro Galazzi, Alessandro Nobili, Mauro Tettamanti, Igor Monti, Alessia Antonella Galbussera, Ernesto Crisafulli, Domenico Girelli, Alessio Maroccia, Daniele Gabbiani, Fabiana Busti, Alice Vianello, Marta Biondan, Filippo Sartori, Paola Faverio, Alberto Pesci, Stefano Zucchetti, Paolo Bonfanti, Marianna Rossi, Ilaria Beretta, Anna Spolti, Sergio Harari, Davide Elia, Roberto Cassandro, Antonella Caminati, Francesco Cipollone, Maria Teresa Guagnano, Damiano D’Ardes, Ilaria Rossi, Francesca Vezzani, Antonio Spanevello, Francesca Cherubino, Dina Visca, Marco Contoli, Alberto Papi, Luca Morandi, Nicholas Battistini, Guido Luigi Moreo, Pasqualina Iannuzzi, Daniele Fumagalli, Sara Leone, Josine A. Oud, Meryem Baysan, Jeanette Wigbers, Lieke J. van Heurn, Susan B. ter Haar, Alexandra G.L. Toppenberg, Laura Heerdink, Annekee A. van IJlzinga Veenstra, Anna M. Eikenboom, Julia Wubbolts, Jonathan Uzorka, Willem Lijferink, Romy Meier, Ingeborg de Jonge, Sesmu M. Arbou, Mark G.J. de Boer, Anske G. van der Bom, Olaf M. Dekkers and Frits Rosendaal.

