## Appendix for "External validation of risk scores to predict in-hospital mortality in patients hospitalized due to coronavirus disease 2019"

**Running head:** Validation of COVID-19 risk scores

Shermarke Hassan^1,2^, Chava L. Ramspek^2^, Barbara Ferrari^3^, Merel van Diepen^2^, Raffaella Rossio^3^, Rachel Knevel^4^, Vincenzo la Mura^1,3^, Andrea Artoni^5^, Ida Martinelli^5^, Alessandra Bandera^1,6^, Alessandro Nobili^7^, Andrea Gori^1,6^, Francesco Blasi^1,8^, Ciro Canetta^9^, Nicola Montano^10^, Frits R. Rosendaal^2^, Flora Peyvandi^1,3^, LUMC‐COVID‐19 Research Group, COVID-19 Network working group

1. Università degli Studi di Milano, Dipartimento di Fisiopatologia medico-chirurgica e dei trapianti, Milan, Italy
2. Leiden University Medical Center, Department of Clinical Epidemiology, Leiden, the Netherlands
3. Fondazione IRCCS Ca' Granda Ospedale Maggiore Policlinico, U.O.C. Medicina Generale Emostasi e Trombosi, Milan, Italy
4. Leiden University Medical Center, Department of Rheumatology, Leiden, the Netherlands
5. Fondazione IRCCS Ca' Granda Ospedale Maggiore Policlinico, Angelo Bianchi Bonomi Hemophilia and Thrombosis Centre, Milan, Italy
6. Fondazione IRCCS Ca' Granda Ospedale Maggiore Policlinico, Infectious Disease Unit, Milan, Italy
7. Istituto di Ricerche Farmacologiche Mario Negri IRCCS, Department of Health Policy, Milan, Italy
8. Fondazione IRCCS Ca' Granda Ospedale Maggiore Policlinico, Respiratory Unit and Cystic Fibrosis Adult Center, Milan, Italy.
9. Fondazione IRCCS Ca' Granda Ospedale Maggiore Policlinico, Department of Medicine, High Care Internal Medicine Unit, Milan, Italy
10. Fondazione IRCCS Ca' Granda Ospedale Maggiore Policlinico, Medicina Generale Immunologia e Allergologia, Milan, Italy.

| **Correspondence address** | **Manuscript information** |
| --- | --- |
| Dr. F. Peyvandi | Word count text: 3654 |
| Università degli Studi di Milano | Figure/table count: 2 tables, 4 figures |
| Department of Pathophysiology and Transplantation | Word count abstract: 249 |
| Via Francesco Sforza 35, 20122, Milan, Italy | Reference count: 36 |
| Tel: +39 0250320288 |  |
| |  |
| **Keywords:** SARS-CoV-2, COVID-19, prediction, mortality | |

**4C mortality score: calculation of original model and recalibrated models**

**Calculating the linear predictor of the original 4C mortality score**

The coefficients of the final logistic regression model are reported in Appendix 5A of the original 4C mortality score publication. (Knight SR, Ho A, Pius R, et al. Risk stratification of patients admitted to hospital with covid-19 using the ISARIC WHO Clinical Characterisation Protocol: Development and validation of the 4C Mortality Score. *BMJ*. 2020;370)

Using this information, we calculated the linear predictor (LP) of the original model using the formula below:

*Table S1*

| **Formula** | **Example*** |
| --- | --- |
| -4.203  + Age = 50-59 * 0.687  + Age = 60-69 * 1.337  + Age = 70-79 * 1.842  + Age ≥ 80 * 2.252  + Male sex * 0.172  + 1 comorbidity * 0.300  + ≥2 comorbidities * 0.532  + Respiratory rate = 20-29 breaths/minute * 0.232  + Respiratory rate ≥ 30 breaths/minute * 0.649  + Oxygen saturation on room air (%) < 92 * 0.577  + Glasgow Coma Scale score < 15 * 0.558  + Blood urea nitrogen (BUN) = 7-14 mmol/L * 0.439  + Blood urea nitrogen (BUN) > 14 mmol/L * 1.011  + CRP concentration = 50-99 mg/dL * 0.363  + CRP concentration ≥100 mg/dL * 0.74 | -4.203  + (0 * 0.687)  + (0 * 1.337)  + (0 * 1.842)  + (1 * 2.252)  + (1 * 0.172)  + (0 * 0.300)  + (1 * 0.532)  + (0 * 0.232)  + (1 * 0.649)  + (1 * 0.577)  + (0 * 0.558)  + (0 * 0.439)  + (0 * 1.011)  + (0 * 0.363)  + (1 * 0.74) |

* Example patient is male, 85 years old, has 3 comorbidities (diabetes, COPD and chronic hypertension). At admission, his respiratory rate was 40 breaths/minute, oxygen saturation on room air was 90%, and he scored the full 15 points on the Glasgow Coma Scale score. Furthermore, blood urea nitrogen (BUN) was 4.5 mmol/L, and CRP was 300 mg/dL. The linear predictor for this patient is 0.719 and predicted probability of mortality is 0.67.

The resulting linear predictor (LP) gives the individual risk of mortality, on the logit (log-odds) scale.

**Standard model**

To obtain the individual predicted probability of mortality, the linear predictor (LP) must be converted from the logit (log-odds) scale to the probability scale. For this, the following formula was used:

*exp(LP) / (1 + exp(LP))*

**Recalibrating the model**

We recalibrated the models, using the methods proposed by Steyerberg (Steyerberg EW. Clinical Prediction Models: A Practical Approach to Development, Validation, and Updating. 2009).

The first way to recalibrate the model was to re-estimate the intercept (*α*). This was done by fitting a new logistic regression model to the data with the following form:

*Logit(y) = α + 1*LP*

The coefficient for the linear predictor is fixed at 1, and the only coefficient in this model that can be freely estimated is the intercept (*α)*. To calculate the individual predicted probability for mortality *y*, the following formula can be used:

*y = exp(α + LP) / (1 + exp(α + LP))*

The second (and more extensive) way to recalibrate the model was to re-estimate both intercept (*α*) and the coefficient (*β_1_*) for the linear predictor:

*Logit(y) = α + β_1_*LP*

To calculate the individual predicted probability for mortality *y*, the following formula can be used:

*y = exp(α + β_1_*LP) / (1 + exp(α + β_1_*LP))*

**Model formulas**

For each fitted model reported in the manuscript, we have reported the corresponding formula to calculate the individual predicted probability are shown below:

*Table S2*

| 4C Model | Formula |
| --- | --- |
| Original model | exp(LP) / (1 + exp(LP)) |
| Lombardy cohort, recalibration method 1 | exp(-0.2984 + LP) / (1 + exp(-0.2984 + LP)) |
| Lombardy cohort, recalibration method 2 | exp(-0.0946 + 1.2954*LP) / (-0.0946 + 1.2954*LP)) |
| Leiden cohort, recalibration method 1 | exp(-1.282 + LP) / (1 + exp(-1.282 + LP)) |
| Leiden cohort, recalibration method 2 | exp(-1.094 + 1.457*LP) / (-1.094 + 1.457*LP)) |

*LP: linear predictor, see table S1 for calculation*
